# Admission rate, indications, maternal outcomes, and factors associated with mortality among obstetric intensive care unit admissions at a tertiary referral hospital in Tanzania

**DOI:** 10.64898/2026.11.10.26367705

**Authors:** Raymond Oyugi Samuel, Godfrey Barabona, Herry Kimwela, Sunil Samji, Wilfred Meza, Muzdalfat Abeid

**Author notes:** Corresponding author Email address (ROS).

## Abstract

**Background:** About 0.07% to 8% of parturients will require intensive care unit (ICU) admission during pregnancy, childbirth or puerperium, with great regional variations. When this occurs, it increases morbidity, mortality, long-term physical and psychological sequelae as well as cost. This study sought to examine a five-year admission rate, indications and outcomes of parturients with severe morbidities admitted in the ICU at a tertiary referral facility and teaching hospital in Tanzania.

**Methodology:** A retrospective cross-sectional study was conducted at Muhimbili National Hospital in Dar es Salaam city that included all women who were admitted to the obstetric ICU between 2019 and 2023. Data were retrospectively collected using a pre-tested checklist from all 1526 obstetric ICU admissions, out of 29391 obstetric unit admissions, during the study period. Sources of data were both case files and electronic registries in the ICU and labour ward. Multivariable logistic regression was used to identify factors associated with maternal mortality, with results presented as adjusted odds ratios (aOR) while indications of ICU admission and maternal outcomes were presented descriptively by using frequencies and tables.

**Results:** Out of 29391 obstetric unit admissions, 1526 required ICU, resulting into obstetric ICU admission rate of 5.2%. Hypertensive disorders were the leading cause of admission (73.7%) followed by obstetric haemorrhages (15%). Frequent ICU interventions included; mechanical ventilation (50%), anticonvulsants (26%), blood transfusion (15%), inotropes (4.2%), and dialysis (3.5%). Maternal fatality rate was 14%. In multivariable regression analysis, vaginal delivery was associated with 47% higher odds of mortality compared to caesarean section (aOR 1.47, 95% CI 1.03-2.11, p=0.036). A moderate ICU stay of 3-4 days showed a 91% protective effect relative to stays of ≤2 days (aOR 0.09, P < 0.001). 5-7day stays were also protective (aOR 0.32, p=0.006). Anticonvulsant use was strongly protective against mortality (aOR 0.02, p < 0.001).

**Conclusion:** This five years review of obstetric ICU admissions (2019-2023) showed that, approximately one in 20 obstetric patients required critical care, with one in seven of ICU admissions resulting in maternal death. Hypertensive disorders of pregnancy accounted for majority of ICU admissions, and deaths occurred predominantly during the early phase of critical care. These findings underscore the need to strengthen antenatal risk identification, optimise referral and emergency obstetric care, ensure timely magnesium sulphate administration, and establish obstetric high dependency units to facilitate earlier interventions. Such measures could reduce the burden on ICUs while improving maternal outcomes.

## Introduction

Majority of pregnancies end uneventfully with a healthy mother and newborn, a few however may end up with unintended outcomes, caused by physiologic and anatomic changes that occur during pregnancy that put stress on different body organ systems limiting their ability to compensate during stressful conditions. These changes heighten risk of complications involving multiple organ systems during pregnancy, labour, delivery, and after delivery [1]. While there is no standardised definition of severe maternal morbidity, some researchers have defined it as an unintended outcome resulting in short-term or long-term consequences to a woman’s health, particularly that which requires admission into the Intensive Care Unit (ICU) for monitoring or treatment, such as eclampsia [2,3].

It is estimated that 0.07% to 8% of women require ICU admission at some point during pregnancy, parturition, or during puerperium, with great geographical and regional variations [4]. More evidence showed there is disparity between low resource settings compared to high income regions. For example, the incidence of obstetric ICU admission in France was 3.6% [5] compared to 15% in India [6]. Similarly, only about 1% of obstetric patients were admitted in ICU in China [7] versus 15.4% of the same category of women in Nigeria [8], and 25.7% in Ghana [9]. Previous studies have linked this to poverty, poor health seeking behavior, limited resources, and lack of access to high-risk obstetric care [10–12].

Significant geographical and regional variations in the indications of obstetric ICU admission have been reported. Some authors found hypertensive disorders of pregnancy the leading cause of ICU admission among women with severe morbidities [4,7–9,13,14], while others have observed higher frequency of severe obstetric haemorrhages, maternal sepsis, and pre-existing cardiac diseases [15–19]. The observed geographic and regional differences in the indications of obstetric ICU admissions indicate unpredictability and complexity of maternal illnesses, especially at the critical stage. Nevertheless, these differences call for local studies to understand the reasons for ICU admission in our setting to design and implement interventions based on the local context.

Maternal mortality rates in many low-income countries have stagnantly been high despite several regional and international commitments, estimated to be 15 times the developed world [14]. However, Tanzania has made substantial progress in reducing maternal deaths gearing towards the Sustainable Development Goals (SDGs) of reducing maternal deaths to lower than 70 per 100,000 live births by 2030. Recent data from the Tanzania Demographic and Health Survey and Malaria Indicator Survey (TDHS-MIS) indicated the country has reduced its maternal mortality ratio by nearly 50% in the span of ten years from 2012 [20]. It is noteworthy that majority of the deaths occur in the critical care settings. Previous evidence from a tertiary referral teaching hospital in Tanzania reported obstetric ICU fatality rate of 20% [21], which is only about half of the ICU mortality rates documented in some low resource settings in the sub-Saharan African countries [19,22,23]. Therefore, for any significant reduction in maternal mortalities to be evident, as well as lowering other associated morbidities and cost of care [24,25], there is urgent need to improve obstetric critical care services. This study aimed to examine the rate of obstetric ICU admission, indications, and maternal outcomes, as well as the associated factors at a tertiary referral and a teaching hospital in Tanzania to provide evidence for informing practice and guiding policy change in obstetric critical care.

## Materials and methods

### Study design and setting

A five-year retrospective cross-sectional study was conducted at Muhimbili National Hospital (MNH) from 1^st^ January 2019 to 31^st^ December 2023, assessing the rate of admission, indications, and outcomes of parturients with severe morbidities admitted to the obstetric critical care unit. MNH is the largest public referral facility in Tanzania, providing tertiary care services in Dar es Salaam city serving over 5.4 million residents (Census 2022) and other referral patients residing in nearby regions. The hospital has over 1500 bed capacity and five specialised ICUs (obstetric, medical, surgical, paediatrics, and neonatal), all adequately equipped to provide critical care services. The obstetric ICU has 15 beds all equipped with mechanical ventilators. It is an open ICU, run by a dedicated team of obstetricians, intensivists, anaesthesiologists, nurses and other allied healthcare providers. Rarely, when adult ICUs are full, female patients may be admitted into the obstetric ICU without obstetric indications. During the study period, there were 1526 admissions to the obstetric critical care, 66 of them were not due to obstetric reasons. The hospital has 18 operating rooms, of which four are dedicated for obstetric and gynaecological surgeries. There are 15 delivery beds in the labour suit where all vaginal births are conducted, except a few sick mothers that deliver in the obstetric ICU.

Following obstetric ICU and labour ward admissions, all patients’ information are documented in the registry books and electronically. Recorded data include name, age, registration number, parity, diagnosis (indication of admission), mode and date of delivery, as well as foetal and maternal outcomes and their dispositions. There were a total of 1526 obstetric ICU admissions between January 2019 and December 2023.

### Sampling

A consecutive (census) sampling approach was used. All obstetric ICU admissions from1^st^ January 2019 to 31^st^ December 2023 were consecutively assessed for eligibility for inclusion. Inclusion criterion was patients who were admitted to the obstetric ICU, while patients with missing information of interest or admitted with non-obstetric reasons were excluded.

### Data collection

A pre-tested checklist was used to extract data from June to December 2024. Research assistants were responsible for data extraction under supervision from the principal investigator who had access to the all-relevant registries that could identify participants during the period of the study but not after. Variables of interest included socio-demographics and obstetric characteristics, indication of ICU admission, date and mode of delivery as well as foetal and maternal outcomes and their disposition.

### Statistical analysis

Of the 1526 obstetric ICU admissions, 211 patients had missing information from either torn pages or incomplete documentation while 66 were admitted with non-obstetric conditions, and therefore excluded. Data of 1249 admissions who met inclusion criteria were analysed using an SPSS ver. 20 (Chicago, Illinois, USA). Descriptive summary of frequencies, percentages and tables, were applied to present the rate of ICU admission, indications of admission, foetal, and maternal outcomes. Univariable and multivariable logistic regression were applied to analyse factors associated with maternal outcomes. Variables with p < 0.2 in the univariable analysis were entered into the multivariable model, and results were reported as crude (cOR) and adjusted odds ratios (aOR) with 95% confidence intervals (CI). Statistical significance was set at p < 0.05.

### Ethical considerations

This study received waiver of consent from the Institutional Review Board (IRB). Ethical clearance was obtained from the IRB of MNH, reference number MNH/TRCU/IRB/PERM/2024/126. Collected data did not include patients’ identity such as names and registration numbers to ensure confidentiality. Only research assistants responsible for data collection and the principal investigator had access to the registries during data collection process. Access to the data during entry and analysis from our computer was password-protected and accessible to the research team only. All the data were completely anonymised with no patient identifiable information. Data will be completely destroyed five years after completion of this study.

## Results

During the study period, there were 29391 total admissions in the obstetric unit, of which 1526 required ICU admission, resulting into an overall ICU admission rate of 5.2%. However, 277 patients did not meet inclusion criteria. Data of 1249 patients were processed and analysed (Fig. 1).

**Figure 1.**
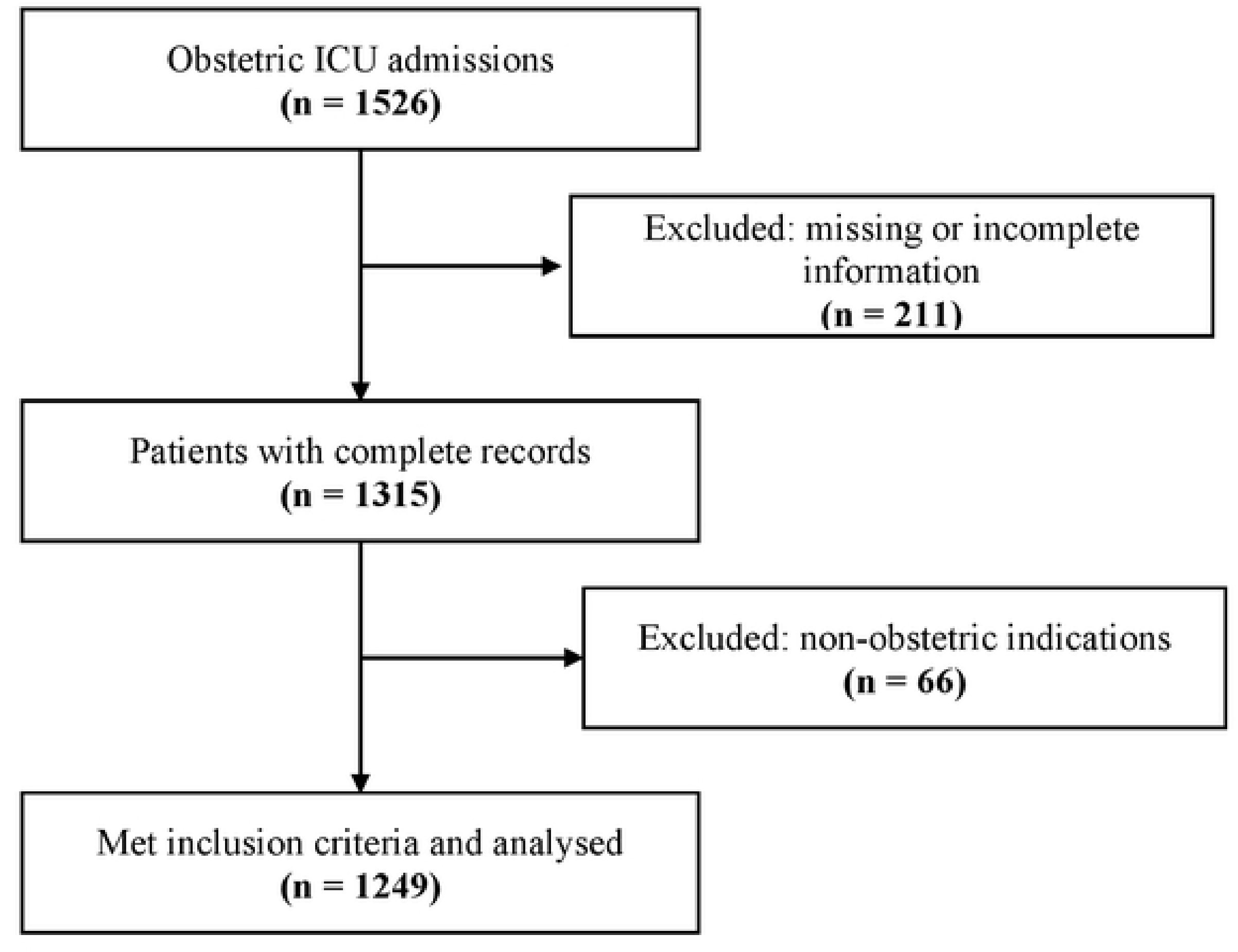
Study population flow diagram.

### Sociodemographic and obstetric characteristics

Majority of women in this study were in the age group of 18-40 (93.5%) followed by those above 40 years (4.3%), while teenage pregnancy constituted 2.2% of all ICU admissions. About 56% of patients underwent caesarean section while 44% delivered vaginally. The median duration of ICU stay was five days. About one third of foetuses had low Apgar scores as well as low birth weights. Nearly 69.7% of the foetuses were healthy, 19.5% required admission into neonatal ICU (NICU), while 11% died (Table 1).

**Table 1.** Sociodemographic, obstetric and clinical characteristics of participants (n = 1249)

| **Variable** |  | **Frequency (n)** | **Percent (%)** |
|---|---|---|---|
| **Maternal age group (years)** |  |  |  |
|  | < 18 | 27 | 2.2 |
|  | 18 – 40 | 1168 | 93.5 |
|  | >40 | 54 | 4.3 |
| **Parity** |  |  |  |
|  | Primigravida | 357 | 28.6 |
|  | Para 1 – 2 | 563 | 45.1 |
|  | Para 3 – 4 | 287 | 23 |
|  | Para 5 and above | 42 | 3.4 |
| **Mode of delivery** |  |  |  |
|  | Vaginal | 552 | 44.2 |
|  | Cesarian section | 697 | 55.8 |
| **Median ICU stay (IQR) (days)** |  | 5 (3, 7) |  |
| **Duration of ICU stay** |  |  |  |
|  | Short ICU stay (≤ 2 days) | 114 | 9.1 |
|  | Moderate ICU stay (3 – 4 days) | 420 | 33.6 |
|  | Prolonged ICU stay (5 – 7 days) | 439 | 35.1 |
|  | Very prolonged ICU stay (> 7 days) | 276 | 22.1 |
| **Foetal clinical characteristics and outcomes** |  |  |  |
| **APGAR score** |  |  |  |
|  | < 7 | 387 | 31 |
|  | 7 – 10 | 862 | 69 |
| **Birth weight** |  |  |  |
|  | < 2.5 kg | 387 | 31 |
|  | ≥ 2.5 kg | 862 | 69 |
| **Foetal disposition** |  |  |  |
|  | NICU | 243 | 19.5 |
|  | Normal ward (ward 36) | 871 | 69.7 |
|  | Died | 135 | 10.8 |

### Percentage distribution of indications of ICU admission

Hypertensive disorders of pregnancy were the leading cause of obstetric ICU admission, largely due to preeclampsia (35.1%), eclampsia (32.8%), and haemolysis, elevated liver enzymes and low platelets (HELLP) syndrome (5.4%), followed by obstetric haemorrhages; postpartum (10.7%) and antepartum (4.2%). Other indications included, Disseminated intravascular coagulopathy (3.5%), maternal sepsis (2.7%), and thromboembolisms/ amniotic fluid embolism (1.4%) (Figure 2).

**Figure 2.**
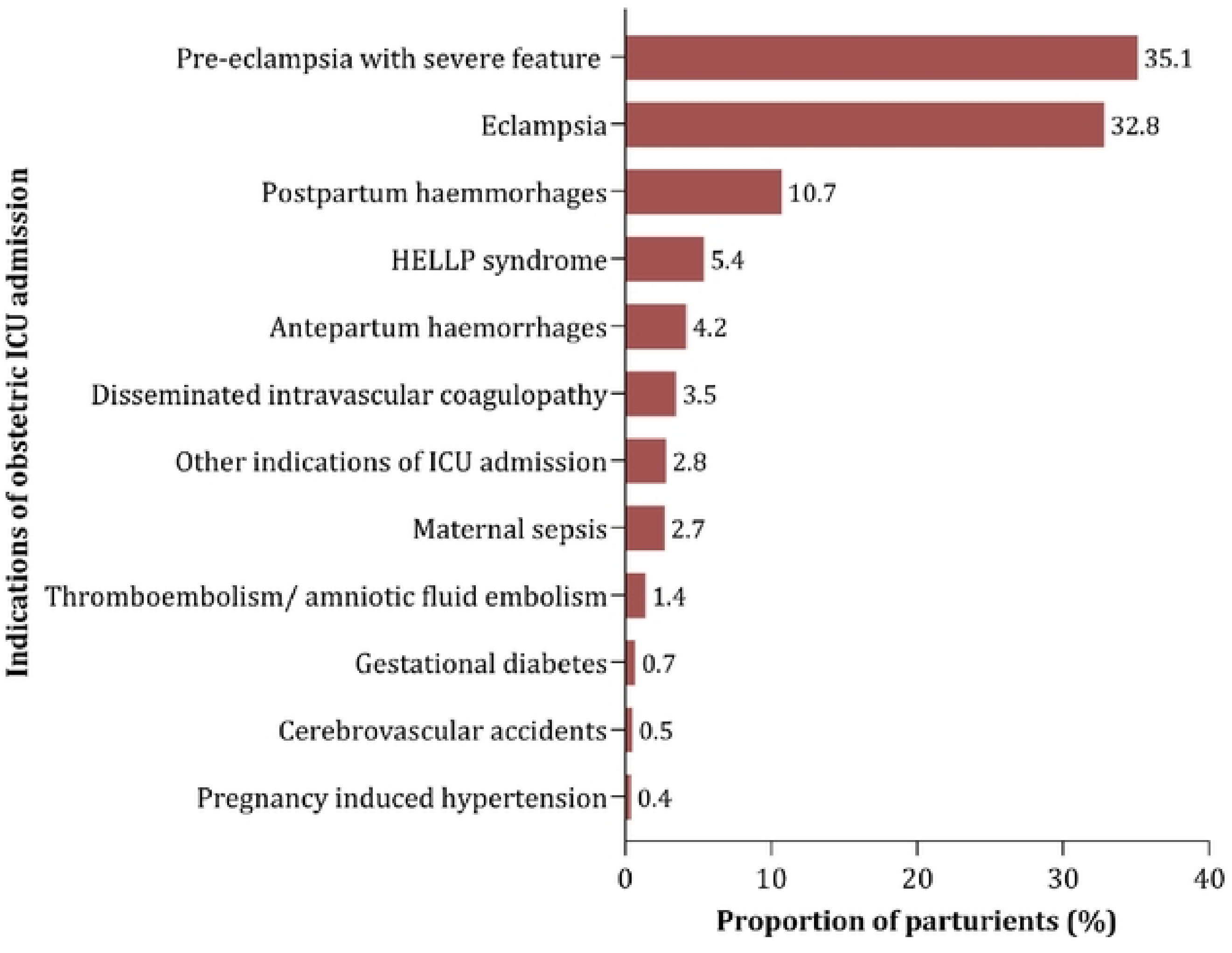
Indications of obstetric ICU admission.

### Percentage distribution of common interventions received in ICU

During ICU stay, 50% of patients required mechanical ventilation, 26% received anticonvulsants for seizure prophylaxis and treatment, 15% were transfused blood and blood products, 4.2% required inotropes or vasopressors, while 3.5% were dialysed (figure 3A)

**Figure 3.**
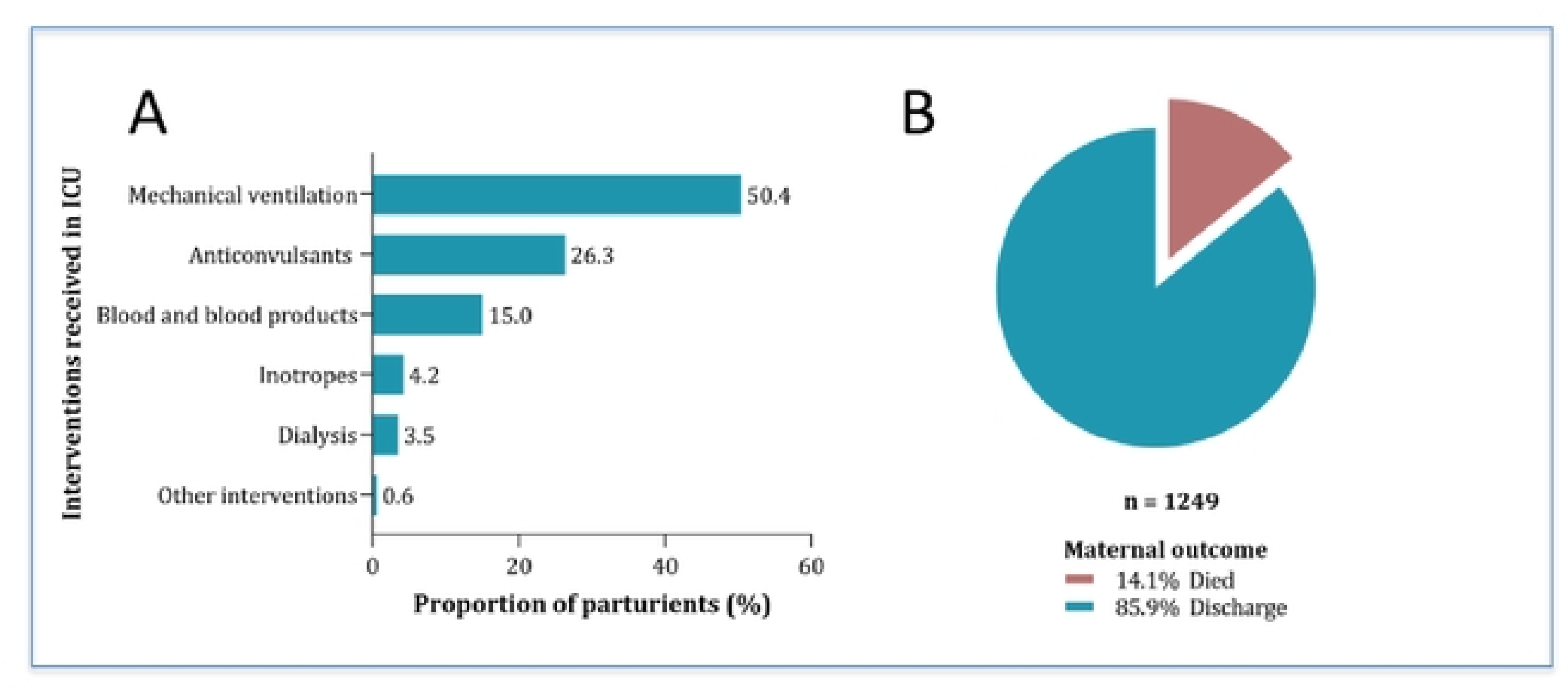
A) Proportions of ICU interventions received in the ICU. B) Pie chart presentation of ICU maternal outcomes.

### Maternal outcomes following ICU admission

Figure 3B presents maternal outcomes in ICU. Out of 1249 ICU admissions, 176 (14%) patients died.

### Univariable and multivariable analysis of factors associated with maternal outcomes in ICU

Table 2 presents univariable and multivariable logistic regression analyses of factors associated with maternal mortality in ICU. Several variables lost significance after adjustment, meaning their crude association was explained by confounding. The following variables retained statistical significance in the multivariable model after adjusting for confounders. Vaginal delivery was associated with 47% higher odds of mortality compared to caesarean section (aOR 1.47, 95% CI 1.03-2.11, p=0.036). A moderate ICU stay of 3-4 days showed a 91% protective effect relative to stays of ≤2 days (aOR 0.09, P < 0.001), while 5-7day stays were also protective (aOR 0.32, p=0.006). Anticonvulsant use was strongly protective (aOR 0.02, p < 0.001).

**Table 2:**
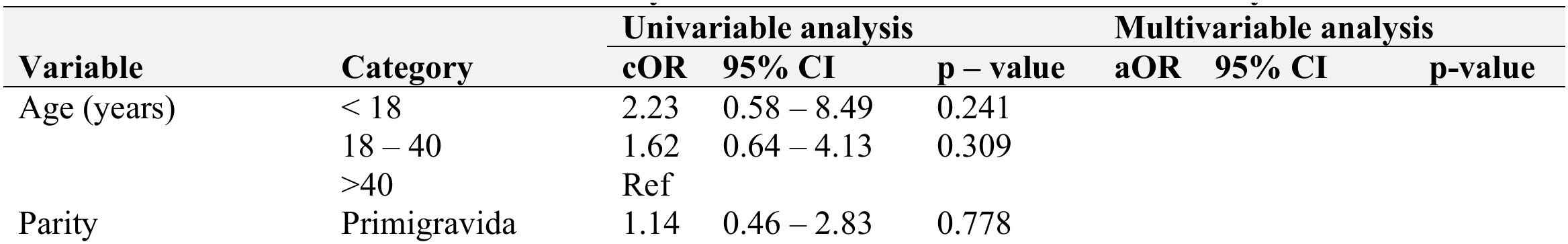
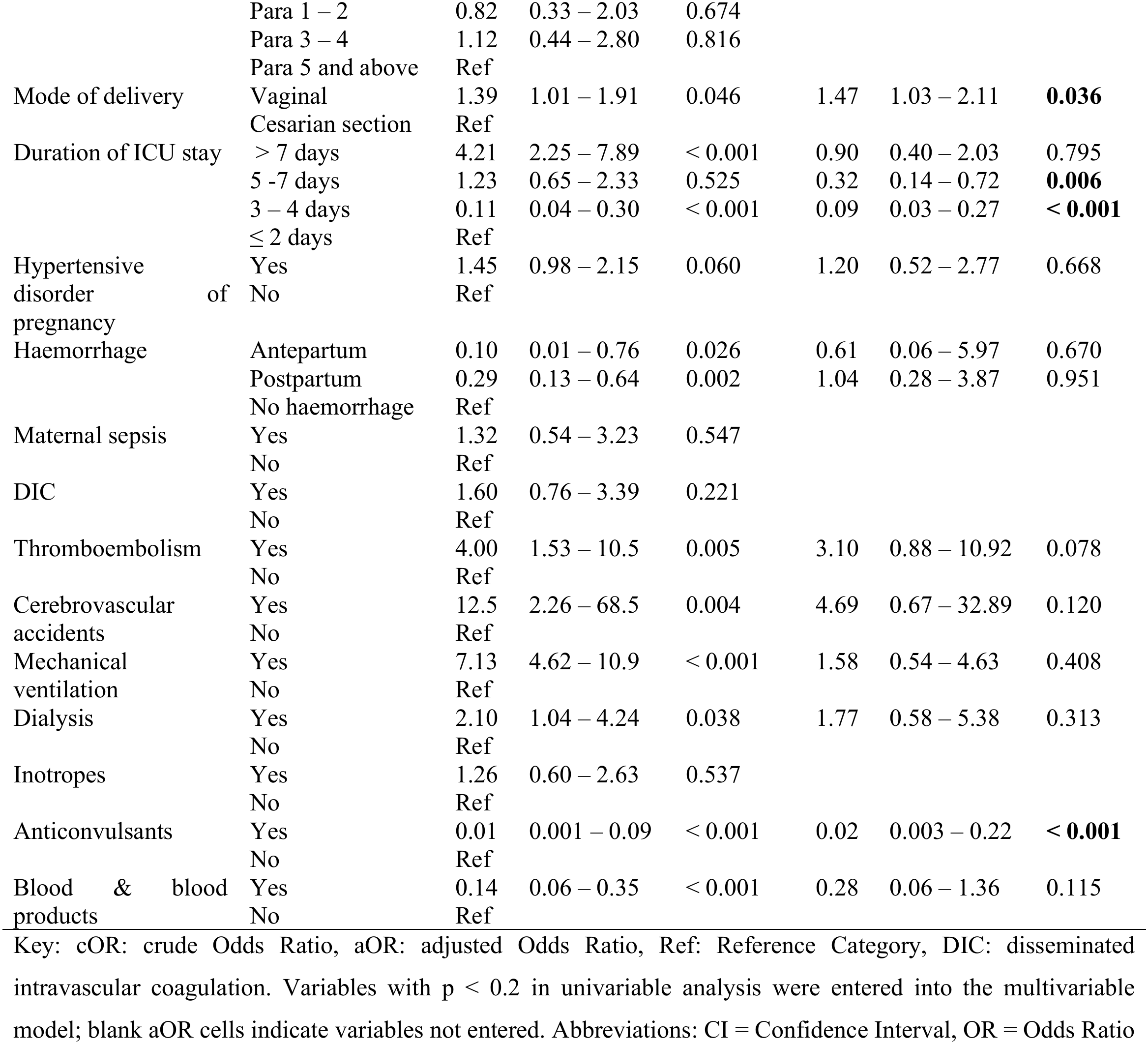
Univariable and multivariable analysis of the factors associated with mortality.

| Variable | Category | Univariable analysis |  |  | Multivariable analysis |  |  |
|---|---|---|---|---|---|---|---|
|  |  | cOR | 95% CI | p – value | aOR | 95% CI | p-value |
| Age (years) | < 18 | 2.23 | 0.58 – 8.49 | 0.241 |  |  |  |
|  | 18 – 40 | 1.62 | 0.64 – 4.13 | 0.309 |  |  |  |
|  | >40 | Ref |  |  |  |  |  |
| Parity | Primigravida | 1.14 | 0.46 – 2.83 | 0.778 |  |  |  |
|  | Para 1 – 2 | 0.82 | 0.33 – 2.03 | 0.674 |  |  |  |
|  | Para 3 – 4 | 1.12 | 0.44 – 2.80 | 0.816 |  |  |  |
|  | Para 5 and above | Ref |  |  |  |  |  |
| Mode of delivery | Vaginal | 1.39 | 1.01 – 1.91 | 0.046 | 1.47 | 1.03 – 2.11 | **0.036** |
|  | Cesarian section | Ref |  |  |  |  |  |
| Duration of ICU stay | > 7 days | 4.21 | 2.25 – 7.89 | < 0.001 | 0.90 | 0.40 – 2.03 | 0.795 |
|  | 5 -7 days | 1.23 | 0.65 – 2.33 | 0.525 | 0.32 | 0.14 – 0.72 | **0.006** |
|  | 3 – 4 days | 0.11 | 0.04 – 0.30 | < 0.001 | 0.09 | 0.03 – 0.27 | **< 0.001** |
|  | ≤ 2 days | Ref |  |  |  |  |  |
| Hypertensive disorder of pregnancy | Yes | 1.45 | 0.98 – 2.15 | 0.060 | 1.20 | 0.52 – 2.77 | 0.668 |
|  | No | Ref |  |  |  |  |  |
| Haemorrhage | Antepartum | 0.10 | 0.01 – 0.76 | 0.026 | 0.61 | 0.06 – 5.97 | 0.670 |
|  | Postpartum | 0.29 | 0.13 – 0.64 | 0.002 | 1.04 | 0.28 – 3.87 | 0.951 |
|  | No haemorrhage | Ref |  |  |  |  |  |
| Maternal sepsis | Yes | 1.32 | 0.54 – 3.23 | 0.547 |  |  |  |
|  | No | Ref |  |  |  |  |  |
| DIC | Yes | 1.60 | 0.76 – 3.39 | 0.221 |  |  |  |
|  | No | Ref |  |  |  |  |  |
| Thromboembolism | Yes | 4.00 | 1.53 – 10.5 | 0.005 | 3.10 | 0.88 – 10.92 | 0.078 |
|  | No | Ref |  |  |  |  |  |
| Cerebrovascular accidents | Yes | 12.5 | 2.26 – 68.5 | 0.004 | 4.69 | 0.67 – 32.89 | 0.120 |
|  | No | Ref |  |  |  |  |  |
| Mechanical ventilation | Yes | 7.13 | 4.62 – 10.9 | < 0.001 | 1.58 | 0.54 – 4.63 | 0.408 |
|  | No | Ref |  |  |  |  |  |
| Dialysis | Yes | 2.10 | 1.04 – 4.24 | 0.038 | 1.77 | 0.58 – 5.38 | 0.313 |
|  | No | Ref |  |  |  |  |  |
| Inotropes | Yes | 1.26 | 0.60 – 2.63 | 0.537 |  |  |  |
|  | No | Ref |  |  |  |  |  |
| Anticonvulsants | Yes | 0.01 | 0.001 – 0.09 | < 0.001 | 0.02 | 0.003 – 0.22 | **< 0.001** |
|  | No | Ref |  |  |  |  |  |
| Blood & blood products | Yes | 0.14 | 0.06 – 0.35 | < 0.001 | 0.28 | 0.06 – 1.36 | 0.115 |
|  | No | Ref |  |  |  |  |  |

Key: cOR: crude Odds Ratio, aOR: adjusted Odds Ratio, Ref: Reference Category, DIC: disseminated intravascular coagulation. Variables with p < 0.2 in univariable analysis were entered into the multivariable model; blank aOR cells indicate variables not entered. Abbreviations: CI = Confidence Interval, OR = Odds Ratio

## DISCUSSION

This study examined the frequency, indications, and maternal outcomes of obstetric ICU admissions, and the factors associated with mortality, at a tertiary referral and teaching hospital in Tanzania. About one in 20 obstetric admissions required intensive care, with hypertensive disorders of pregnancy accounting for nearly three-quarters of these, followed by obstetric haemorrhage. Approximately one in seven women admitted to the ICU died. Vaginal delivery was associated with higher odds of mortality compared to caesarean section, whereas receipt of anticonvulsant therapy was independently associated with a reduced risk of death. In addition, an ICU stay of three to four days was associated with lower odds of mortality compared with a stay of two days or less, suggesting that the highest mortality occurred during the early phase of critical illness.

The obstetric ICU admission rate of 5.2% in the present study is high and consistent with the marked global disparities previously reported in critical care admission following delivery. For example, an observational study in China reported a peripartum ICU admission rate of about 1% [26], considerably lower than our finding and those from other sub-Saharan African settings [22,27,28]. However, most of these were single-centre observational studies that used different denominators (deliveries, obstetric admissions or all ICU admissions), which limit direct comparison. At the regional level, a systematic review and meta-analysis from Africa found that obstetric patients accounted for about 17% of all ICU admissions [29]. These disparities likely reflect regional differences in health-system infrastructure, severity of illness at presentation, and access to timely intervention. In low-resource settings, the three delays (in deciding to seek care, reaching a facility, and receiving adequate treatment) have consistently been linked to the escalation of obstetric complications into critical illness [28,29]. As MNH is a tertiary referral hospital, delays at any of these levels may have contributed to its high ICU admission rate. Structural constraints may also contribute. Studies from the African region show that, in the absence of step-down facilities such as HDUs, health facilities often admit even patients who require only observation and monitoring to the ICU, inflating ICU admission rates [27,29]. This is supported by evidence from China, where the introduction of an obstetric HDU substantially reduced ICU admissions [26]. Because MNH had no obstetric HDU during the study period, the lack of this intermediate level of care may partly explain the high ICU admission rate observed.

In the present study, hypertensive disorders of pregnancy were the leading indication for ICU admission (73.7%), followed by severe obstetric haemorrhage (15.0%). This pattern is consistent with earlier findings at MNH [21] and with reports from other settings in Africa and Asia [4,7–9,13,14]. Although the indications for obstetric ICU admission are broadly similar in high-income countries (HICs) and low- and middle-income countries (LMICs), hypertensive disorders account for a far higher proportion of admissions in LMICs (43–70%) than in HICs [27,29,30]. In HICs, a growing share of admissions is due to indirect obstetric causes such as cardiomyopathy, pulmonary hypertension and complications of assisted reproduction [31,32], reflecting older maternal age and more comorbidities. Because hypertensive disorders are largely detectable through routine antenatal screening, their predominance in our cohort suggests gaps in antenatal care (ANC), particularly in early screening, detection and optimisation of medical conditions, and timely referral for specialised care. This is supported by a recent analysis of Tanzanian national survey data, which found that nearly half of pregnant women had an inadequate number of ANC visits [33]. Similarly, at a Nigerian teaching hospital, about 70% of women admitted to the obstetric ICU had not booked for ANC [28]. Strengthening ANC to ensure early detection and optimisation of maternal medical conditions, with timely referral, is therefore essential [34].

The maternal ICU mortality of 14% in our study is high and consistent with a systematic review and meta-analysis from the African region [29] and with observational studies from other low-resource settings in Africa and Asia [19,28,35]. In contrast, studies from some European countries have reported obstetric ICU mortality as low as 2% [18]. Consistent with this, a systematic review found obstetric ICU mortality to be nine times higher in LMICs than in HICs [36]. These marked regional differences in maternal ICU mortality highlight an opportunity for improving maternal critical care outcomes in LMICs, given that about 50% of maternal deaths are considered avoidable [14,37]. Although this study did not assess the reasons for the high ICU mortality in our setting, studies from other resource-limited settings have reported infrastructural constraints, late presentation, and inadequate access to ANC as drivers of high fatality rates [26–29,35]. A study comparing obstetric critical care outcomes between American and Indian public hospitals found that only 27% of Indian women (versus 86% of American women) had attended ANC, and that mortality was significantly lower among patients who had attended regular ANC than among those who had not [38]. Despite that study’s limitations, the role of ANC in overall maternal health is difficult to ignore. Future studies at MNH should examine how these factors directly or indirectly influence obstetric ICU outcomes.

In the multivariable analysis of factors associated with maternal ICU outcomes, only three variables remained statistically significant after adjustment for confounders. An ICU stay of three to seven days was significantly associated with survival compared with a stay of two days or less, likely reflecting late presentation and rapid clinical deterioration before adequate treatment could be initiated. A similar study in Asia reported a mean ICU stay of 5 days and linked stays of one day to mortality, attributing this to late referral and poor ANC; 60% of its participants had not attended ANC [39]. Other studies in the African region found that patients who stayed in the ICU for less than five days were two to three times more likely to die than those with longer stays [40,41], findings attributed to poor referral systems, shortages of essential supplies, medicines, human resource, and late presentation. How these factors influenced ICU stay and mortality in our setting was beyond the scope of this study.

In the present study, vaginal delivery was associated with 47% higher odds of mortality compared with caesarean section (aOR 1.47, 95% CI 1.03-2.11, p=0.036). This finding may reflect earlier clinical intervention among women who underwent caesarean section. Previous studies have noted that critical illness may necessitate urgent operative delivery to halt ongoing pathology, whereas vaginal birth may take longer, particularly in women who are not in labour [42,43]. In contrast, a longitudinal case series study from East Africa found no association between mode of delivery and maternal outcomes among women with eclampsia, although caesarean delivery was significantly associated with poor perinatal outcomes [44]. A larger population-based multicentre study also reported findings contrary to ours, showing that caesarean delivery was an independent predictor of maternal mortality rather than a protective intervention, except in breech presentation, where it independently reduced the overall risk [45]. Similarly, a 10-year case-control study using a nationwide inpatient sample from an HIC found caesarean delivery to be associated with a more than threefold increase in the adjusted odds of maternal death, which rose to about sixfold among primary caesarean deliveries [46]. These discrepancies may reflect differences in study design and setting, patient characteristics and illness severity at presentation, as well as ICU organisation and services between HICs and LMICs.

In addition, anticonvulsant therapy (magnesium sulphate) was strongly protective against ICU mortality (aOR 0.02, p < 0.001). Magnesium sulphate is the anticonvulsant of choice for the prevention and treatment of eclamptic seizures, despite debate about its anticonvulsant efficacy [47–49]. A four-year retrospective review of women admitted to the ICU with complications of hypertensive disorders of pregnancy at a tertiary hospital in South Africa found that failure to administer magnesium sulphate when indicated was a significant predictor of maternal mortality [48]. Furthermore, a narrative review reported that prophylactic magnesium sulphate more than halved the risk of eclampsia compared with placebo [50], underscoring its central role in both the prevention and treatment of eclamptic seizures.

### Study strengths and limitation

This study has several strengths. It aligns with local and global efforts towards the Sustainable Development Goals (SDGs) by identifying measures to reduce maternal mortality, particularly among critically ill women in the ICU. It also highlights important outcomes of obstetric ICU admission, providing a basis for planned future projects and for practice and policy changes aimed at improving early recognition of warning signs, timely referral for specialised care, and prompt management of critically ill mothers. This study also has some limitations. First, 211 patients were excluded because key variables were missing or incomplete, owing to torn registry pages and incomplete documentation. Nevertheless, the five-year study period allowed an adequate sample size to be obtained. In addition, patient records are now stored electronically, which should improve data quality for future studies. Second, the retrospective design made the study dependent on the accuracy and completeness of routine records; however, multiple data sources were used to reduce misclassification of indications and outcomes arising from inconsistent documentation. Like other retrospective data, these records were also prone to bias and confounding. However, the use of multiple clinical registries and data sources (registry books and electronic records) improved completeness and reduced selection bias. In addition, multivariable logistic regression was used to adjust for confounders. Third, this was a single-centre study conducted at the country’s highest-level referral facility, which serves a high-risk population, and may therefore not be representative of other facilities. However, our findings may be applicable to other tertiary referral facilities, depending on their institutional protocols and local resource constraints. Finally, because of the cross-sectional design, only associations could be described; causation could not be established.

## Conclusion

This study found that approximately one in twenty obstetric admissions required intensive care (5.2%), with an obstetric ICU mortality rate of 14%. Hypertensive disorders of pregnancy were the leading indication for ICU admission, followed by obstetric haemorrhage. Caesarean delivery, anticonvulsant therapy, and an ICU stay of three to seven days (compared with two days or less) were independently associated with lower odds of maternal death. The predominance of largely preventable hypertensive complications, together with the concentration of deaths within the first two days of ICU admission, highlight gaps in antenatal risk identification, timely recognition of clinical deterioration, referral pathways, and emergency obstetric management. Strengthening antenatal screening for hypertensive disorders, ensuring timely administration of magnesium sulphate, improving referral systems, and establishing an obstetric high dependency unit could reduce the need for intensive care and improve maternal survival. Prospective multicentre studies are warranted to identify the modifiable determinants of early ICU mortality and to evaluate interventions aimed at improving outcomes among critically ill obstetric patients.

## Data Availability

Data set is available in the supporting information section

## Author contributions

ROS conceptualised, designed the study, supervised data collection and analysis and drafted the first manuscript. GB, HK, SS, WM, and MA interpreted results.

All authors read the first manuscript and approved the final document.

## Acknowledgement

Not applicable

